# Prevalence and Determinants of Double and Triple Burden of Malnutrition Among School Going Children and Adolescents in Zanzibar, 2022

**DOI:** 10.1101/2025.02.28.25323109

**Authors:** Esther Ngadaya, Anna Mosses, Germana Leyna, David Solomon, Hawa Msola, Fatma Ally Said, Hope Masanja, Gibson Kagaruki, Ramadhani Mwiru, Asha Salmin, Kahabi Isangula, Miriam Kiungai, Kombo Mdachi Kombo, Geofrey Mchau, Joyce Ngegba, Patrick Codjia

## Abstract

**Background:** Malnutrition stands as a profound global health concern, and its dimensions are evolving. In Zanzibar, the burden of malnutrition especially among school going children is not well unknown, as such, this study was conducted to assess the prevalence and factors associated with double and triple burden of malnutrition among school going children and adolescents in Zanzibar.

**Methods:** This was a School-based cross-sectional study involving mainly quantitative data collection method. Data was collected as part of the National School Health and Nutrition Survey of 2022, which included primary and secondary school children and adolescents aged 5-19 years, from Zanzibar. Anthropometric measurements and hemoglobin levels of selected students were collected. A multinomial regression model was used to assess factors associated with double burden of malnutrition (DBM) and the triple burden of malnutrition (TBM). Z-scores for weight, height and body mass index for the scholars aged 5-19 years were generated using WHO AnthroPlus and data analysis was done using Stata Version 17.

**Results:** A total of 2556 primary and secondary school children were enrolled, 51.7% (n= 1,322) were girls. Almost 2 in 5 were individuals with 10-14 years, and most of them were from primary schools. Slightly over 5 in 10 were residing in urban areas. Overall, the prevalence of malnutrition defined as malnutrition of any kind (stunting or underweight/thinness or overweight or anemia) in Zanzibar was 58.4 per cent. The overall prevalence of DBM defined by the coexistence of both undernutrition and over-malnutrition in Zanzibar was 12.0 per cent. Similarly, the overall prevalence of TBM defined as the coexistence of undernutrition, anemia, and overnutrition in Zanzibar was 1.8%. In an adjusted multinomial regression model, the prevalence of DBM was 30% lower if the child was 5 to 9 years 0.7 (95% CI, 0.5–0.9), p = 0.02), and 1.5-fold greater if the student was living in a lowest wealth quantile family (1.5(95%CI, 1.01-2.3), p=0.04). In the contrary, the prevalence of single malnutrition was 1.4-fold greater if the student was a girl (1.4(95%CI,1.2-1.6); p=0.001). Proportion of students with TBM was 2.0-fold greater if there were no school deworming education (2.0(95%CI, 1.0-3.9); p=0.05

**Conclusion:** Over half of the students in Zanzibar are malnourished, with a significant burden of DBM, indicating the need for stringent actions to reduce the prevalence of malnutrition in the country.

## INTRODUCTION

Malnutrition stands as a profound global health concern, and its dimensions are evolving. Classically, malnutrition evoked images of undernourishment and micronutrient deficiencies. Today, it also encompasses the ballooning epidemic of overweight and obesity(1). When undernutrition and overweight/obesity coexist within a population, it’s termed the “double burden of malnutrition”. When one also considers the scourge of micronutrient deficiencies, this escalates to a “triple burden”(2) larmingly, this multifaceted problem is increasingly prominent in low and middle-income countries (3).

Zanzibar, a significant archipelago within Tanzania, finds itself at this complex crossroads. Amidst laudable efforts to combat traditional malnutrition, a new challenge has emerged among its youth: the double and sometimes triple burden, especially evident among students. A 2016 report elucidated that Zanzibar’s obesity rates in school-aged children were eclipsing those of Mainland Tanzania (4). Yet, simultaneously, pockets of undernutrition remain, highlighting the duality of Zanzibar’s malnutrition conundrum.

Globally, the magnitude is daunting. The World Health Organization (WHO) reported in 2019 that over 340 million children and adolescents aged 5-19 were overweight or obese(5). Furthermore, a staggering two billion adults are overweight, with over 650 million suffering from obesity(5). By 2030, the global overweight populace could soar to 2.16 billion. At least 50% of children worldwide ages 6 months to 5 years suffer from one or more vitamin or mineral deficiency, which is linked to the increases the risk of anemia, blindness, measles, diarrhea, and rickets.(6) Such vast numbers carry with them elevated risks for numerous ailments, notably diabetes, cardiovascular disorders, and some types of cancers(1,5)

The 2015-16 Demographic and Health Survey (DHS) for Tanzania revealed that 32% of children under five years were stunted. In contrast, 5% were wasted, and 4% were overweight(7). Zanzibar’s stunting rates ranged from 20-24% (7). However, Zanzibar reported a higher prevalence of overweight and obesity, especially among women of reproductive age, compared to the mainland(4). Furthermore, Tanzania National Nutrition Survey of 2018 showed that the prevalence of overweight in under-fives is 2.8% nationally while in Zanzibar is 2.1%. In addition, in women of reproductive age obesity is 31.7% national while in Zanzibar is 41.8% respectful.

Previous literature recognized the basic and underlying causes of malnutrition(8–10), with few interconnected factors catalyzing the double and triple burden of malnutrition. These range from maternal, child, household and community-level factors socio-economic dynamics, urbanization influxes, global dietary shifts, to sedentary lifestyles facilitated by technological advancements

(3). The rapid urban transition, adoption of Western dietary habits, and declining physical activities among students serve as major contributors in Zanzibar (4). However, few studies if any have examined the double and triple burden of Malnutrition. Therefore, it is necessary to understand the existence of the dual and triple burden of undernutrition among children and adolescents aged 5 to 19 years in Zanzibar. Given the persistent and high levels of the problem in Zanzibar, there is a strong need for action to inform the policy. This study was conducted as part of the National School Health and Nutrition Survey (SHNS) 2022 to assess the prevalence of malnutrition including double and triple burden of malnutrition among school-going children and adolescents aged 5 to 19 years in Zanzibar.

## MATERIAL AND METHODS

### Study design

The study was a school-based cross-sectional study. Data were gathered from students aged between 5 and 19 years who were enrolled in primary and secondary schools during the academic year 2022. Students with serious illnesses or who were unwilling to participate were excluded.

### Study setting

The study was conducted in Zanzibar. Zanzibar is an insular semi-autonomous province which united with Tanganyika in 1964 to form the United Republic of Tanzania with a total population of 1,889,773 persons in the 2022 Census(11). Over one-third of the population is between 5 to 19 years old. Zanzibar is an archipelago in the Indian Ocean, 25–50 km (16–31 mi) off the coast of the African mainland, and consists of many small islands and two large ones: Unguja (the main island, referred to informally as Zanzibar) and Pemba Island. The capital is Zanzibar City, located on the island of Unguja. Zanzibar has a total of 5 regions (3 in Unguja and 2 in Pemba). Almost half of the population (893,169; 47.3%) is found in only one region Mjini Magharibi which is in Unguja. Zanzibar’s marine ecosystem is an important part of the economy for fishing and algaculture and contains important marine ecosystems that act as fish nurseries for Indian Ocean fish populations, and also the main source of protein for its citizens including children and adolescents.

### Sampling technique

We utilized a stratified multi-stage sample design (12). The initial stage involved selecting a sample of schools as the primary sampling unit (PSU) using a probability proportional to size (PPS) method. In the second stage, one class level was selected using a simple random sampling approach (SRS), followed by the selection of individuals from these classes. Each selected class had 28 students evenly distributed by sex (14 girls and 14 boys), chosen using a systematic sampling technique.

### Sample size calculation

The sample was selected to provide a plausible estimate of anthropometric and Hemoglobin levels across different domains, such as national, regional and district levels as well as urban/rural areas, and according to sex. The sample size was estimated at the district level (domains) and aggregated to obtain the regional and national sample. The prevalence of anemia was the preferred indicator, utilizing an indicative value of 50%, with a margin error of 0.5, a significance level of 5%, and a design effect of 1.5.

### Data and Sample collection

#### Social-demographic and economic information

Pre-tested structured questionnaires were used to collect data on demographic characteristics and socioeconomic factors after obtaining a written informed consent form through face-to-face interviews. Questionnaires were developed in English, translated into Swahili and then back translated to English to verify accuracy. Independent variables such as household wealth index was assessed as an indicator of socio-economic status according to a standard approach in equity analysis. Durable household assets indicative of wealth (i.e., telephone, radio, TV, refrigerator, lantern, cupboard, houses with electricity, motorcycle, bicycle, cart etc.) were recorded as (1) “available and in working condition” or (0) “not available and/or not in working condition.” These assets were analyzed using principal components analysis (PCA). The component resulting from this analysis was used to categorize households into two approximate quintiles with the following categories: Lowest, Second, Middle, Fourth and Highest quintile.

#### Anthropometric assessments

Indicators of the nutritional status of children were calculated using standards published by the World Health Organization(13). Anthropometric data were collected by recording the age, weight, and height of the participants, and measurements were done on the school premises. A portable weight scale and a stadiometer with a sliding headpiece were used to measure weight (to the nearest 0.1 kg) and height (to the nearest 0.1 cm), respectively. Each child was weighed with minimum clothing and barefoot. The weighing scale was calibrated using the standard calibration weight of 2 kg iron bars. Height was measured in Frankfurt position (head, shoulder, buttocks, knee, and heels touching the vertical board) to the nearest 0.1 cm. Measurements of weight and height were taken twice and the average was recorded. Analysis for anthropometric variables were performed using WHO Anthro Plus (version 1.0.4). Then anthropometric measurements were converted into anthropometric indices such Height-for-age Z scores (HAZ), Body Mass Index-for-age Z scores (BAZ) and Weight-for-age Z scores (WAZ) based on study objectives. Children whose height-for-age Z-score was below minus two standard deviations (–2 SD) from the median of the WHO reference population were considered short for their age (stunted), children with Body Mass Index (BMI)-for-Age Z scores (BAZ) below –2 standard deviation (SD) of the WHO standard were classified as thin, and those with BAZ above +3 were classified as overweight. Weight-for-age (WAZ)-Children with weight-for-age below minus two standard deviations were classified as underweight.

#### Dietary assessment

For the purpose of this survey 21 items, Food Frequency Questionnaire (FFQ) of foods consumed over the past 7 days, was collected and used to estimate the quality of student’s diets. This FFQ will be referred to as a short FFQ throughout the rest of this manuscript. The short form FFQ was pilot-tested with students. Twenty-one foods items were listed and the students in Zanzibar schools were asked to respond whether they eat/drink the items, or did not eat/drink the items at all, or ate food/drink items once, more than once or don’t know, over the past 7 days from the day of the survey. The data which were obtained in FFQ questionnaire was then used to determine the dietary quality of Zanzibar school students using Prime Dietary Quality Score (PDQS) (1) as follow below.

#### Prime Dietary Quality Score

An un-weighted PDQS (1) was constructed from the short FFQ. Foods were deemed health or unhealthy based on the current WHO guidelines and guided by recommended Daily Allowance (RDA) for school age children and adolescents 5-19 years worldwide. The Prime Diet Quality Score (PDQS) was recently developed using a modified Prime Screen questionnaire as a way to characterize diet quality globally and was associated with a lower risk of coronary heart diseases (CHD) to a large population in USA. PDQS contained 21 food groups; 13 were categorized into healthy, and 8 as unhealthy food groups. Food groups were queried on a 5-point Likert scale: 0= never, 1= once in a week, 2-4 times a week, 5-6 times a week and every day. Points were assigned for consumption of healthy food groups as follows: 0–1 serving/week, 0 points; 2–3 servings/week, 1 point; and ≥4 servings/week, 2 points. Scoring for unhealthy food groups was assigned as follows: 0–1 serving/week, 2 points; 2–3 servings/week, 1 point; and ≥4 servings/week, 0 points. Individual scores were aggregated to generate an overall score which ranged from 0 to 42 possible scores, the higher the score the quality of diet consumed by a respective children or adolescent. The score was also divided into quintiles, quintile 3 indicates the poorest dietary quality (Low), quartile 2 indicates medium dietary quality and quintile 1 indicates the highest in the categorical format.

#### Ethical considerations

Ethical clearance was secured from the Zanzibar Medical Research and Ethics Committee. Consent was obtained from schools and parents/guardians, while assent was sought from participants aged 5-19 years. Consent forms, provided in both English and Kiswahili, detailed the study’s procedures, potential risks and benefits, confidentiality measures, and contact details for the study coordinators. Participants were not financially compensated, but they received a juice drink and biscuits post-blood collection and promptly received their laboratory results and anthropometric measurements.

## RESULTS

### Demographic characteristics

The study was implemented in all 11 districts of Zanzibar. The demographic and socio-economic characteristics of the survey population including age, sex, place of residence, caregiver’s schooling and household wealth status are presented in Table 1 (below). A Slightly higher proportion (51.7%; 1,322) of the study participants were girls. Almost 2 in 5 were individuals from 10-14 years, and most of them were from primary school. Slightly over 5 in 10 were residing in urban areas.

**Table 1:** Social demographic characteristics of the study participants (5-19 years; N=2,556).

| Background Characteristics | Number | Percentage (%) |
| --- | --- | --- |
| **Sex** |  |  |
| Male | 1,234 | 48.3 |
| Female | 1,322 | 51.7 |
| **Age** |  |  |
| 5 – 9 | 784 | 30.7 |
| 10 – 14 | 1,058 | 41.4 |
| 15 – 19 | 714 | 27.9 |
| **Level of Education** |  |  |
| Primary | 1,768 | 69.1 |
| Secondary | 788 | 30.8 |
| **School ownership** |  |  |
| Public | 2463 | 96.4 |
| Private | 93 | 3.6 |
| **Area** |  |  |
| Rural | 1,194 | 46.7 |
| Urban | 1,362 | 53.3 |
| **Wealth quintile** |  |  |
| Lowest | 530 | 20.7 |
| Second | 493 | 19.3 |
| Middle | 513 | 20.1 |
| Fourth | 538 | 21.0 |
| Highest | 482 | 18.9 |
| **District** |  |  |
| Kaskazini A | 184 | 7.2 |
| Kaskazini B | 196 | 7.7 |
| Kati | 196 | 7.7 |
| Kusini | 196 | 7.7 |
| Mjini | 264 | 10.3 |
| Magharibi A | 305 | 11.9 |
| Magharibi B | 332 | 13.0 |
| Wete | 251 | 9.8 |
| Micheweni | 195 | 7.6 |
| Chakechake | 219 | 8.6 |
| Mkoani | 218 | 8.5 |
| **Region** |  |  |
| Kaskazini Unguja | 380 | 14.9 |
| Kusini Unguja | 392 | 15.3 |
| Mjini Magharibi | 901 | 35.3 |
| Kaskazini Pemba | 446 | 17.4 |
| Kusini Pemba | 437 | 17.1 |

### Prevalence of Malnutrition stratified by the type of malnutrition

Figure 1 presents the nutritional status of children and adolescents aged 5 to 19 years stratified by the type of malnutrition and demographic characteristics. Overall, 45.7%, 15.6%, 13.9% and 8.4% respectively were anaemic, stunted, underweight, and overweight/obese.

**Figure 1:**
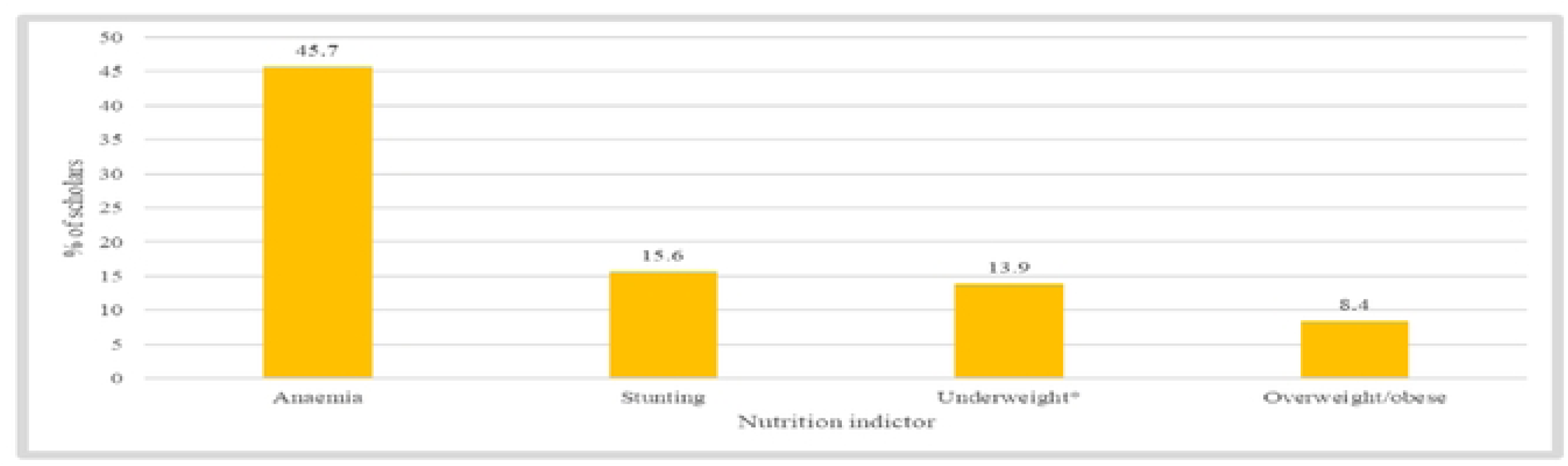
Prevalence of stunting, underweight, anemia and overweight/obese among children and adolescents aged 5 ta 19 years using WHO AnthroPlus Software (n=2556)

### The overall prevalence of malnutrition

Overall, the prevalence of malnutrition defined as malnutrition of any kind (stunting or underweight/thinness or overweight or anaemia) in Zanzibar was 58.4 per cent (Figure 2). At least 44.6 per cent of children had one form of malnutrition.

**Figure 2:**
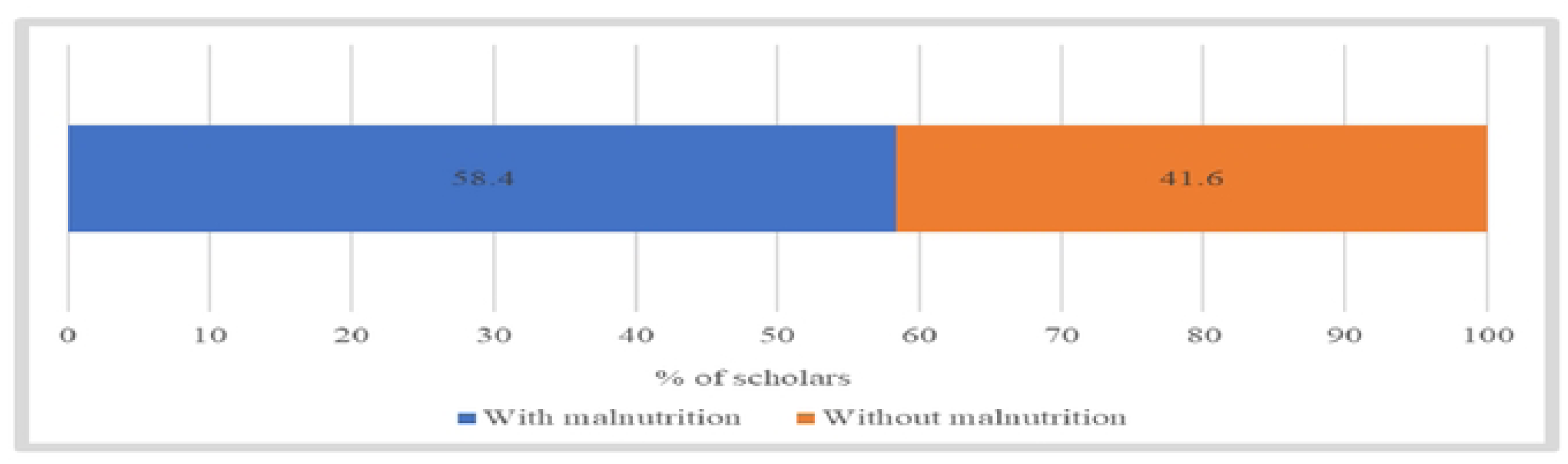
Proportion of children and adolescents aged 5 to 19 years with and without malnutrition in Zanzibar (n=2556).

### Prevalence of Multiple forms of malnutrition

The overall prevalence of double Burden of Malnutrition (DBM) defined by the coexistence of both undernutrition and over-malnutrition in Zanzibar was 12.0 per cent. Similarly, the overall prevalence of the triple burden of malnutrition defined as the coexistence of undernutrition, anemia, and overnutrition in Zanzibar was 1.8% (Figure 3).

**Figure 3:**
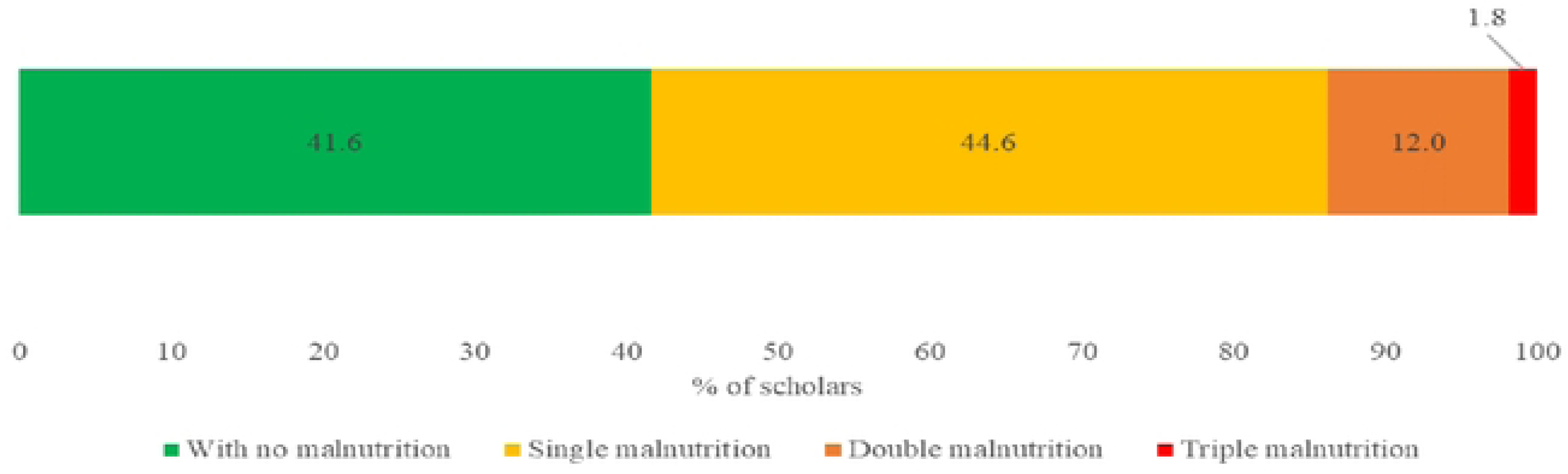
Proportion of children and adolescents aged 5 to 19 years stratified by single and multiple malnutrition; n=2556.

### Burden of malnutrition by socio-economic and demographic characteristics

Table 2 below describe distribution of burden of malnutrition by socio-economic and demographic characteristics. Overall, while the prevalence of single malnutrition was evenly distributed by age, the burden of triple malnutrition was more among 5 to 9 years children. There was no gender variation in the proportion of individuals with triple malnutrition. Government and primary schools’ students and those residing in Magharibi A had the highest proportion with triple malnutrition. The pupils coming from the lowest wealth quintile had a slightly high proportion of children with double and triple burden of malnutrition respectfully

**Table 2:** Distribution of nutrition status by socio-economic and demographic characteristics.

| Variable | n | Single malnutrition (%) | Double malnutrition (%) | Triple malnutrition (%) |
| --- | --- | --- | --- | --- |
| **Age in years** |  |  |  |  |
| 5 to 9 | 784 | 46.2 | 8.6 | 5.2 |
| 10 to 14 | 1058 | 44.2 | 13.2 | 0.0 |
| 15-19 | 714 | 43.6 | 14.0 | 0.7 |
| **Gender** |  |  |  |  |
| Male | 1234 | 40.4 | 13.3 | 1.9 |
| Female | 1322 | 48.5 | 10.8 | 1.7 |
| **Ownership** |  |  |  |  |
| Government | 2463 | 44.9 | 12.0 | 1.8 |
| Private | 93 | 36.6 | 11.8 | 1.1 |
| **Level** |  |  |  |  |
| Primary | 1763 | 44.7 | 11.5 | 2.3 |
| Secondary | 788 | 44.4 | 13.2 | 0.6 |
| **Settings** |  |  |  |  |
| Rural | 1194 | 45.2 | 12.1 | 1.8 |
| Urban | 1362 | 44.1 | 11.9 | 1.8 |
| **District** |  |  |  |  |
| Kaskazini A | 184 | 48.9 | 13.0 | 2.2 |
| Kaskazini B | 196 | 48.0 | 6.6 | 1.5 |
| Kati | 196 | 46.4 | 16.4 | 0.5 |
| Kusini | 196 | 47.5 | 10.7 | 2.5 |
| Mjini | 264 | 40.5 | 10.6 | 1.2 |
| Magharibi A | 305 | 44.6 | 9.8 | 3.0 |
| Magharibi B | 332 | 45.2 | 9.0 | 1.5 |
| Wete | 251 | 46.2 | 14.7 | 2.4 |
| Micheweni | 195 | 40.0 | 11.8 | 0.5 |
| Chakechake | 219 | 42.0 | 14.6 | 1.8 |
| Mkoani | 218 | 42.7 | 17.0 | 2.3 |
| **Region** |  |  |  |  |
| Kaskazini Unguja | 380 | 48.4 | 9.7 | 1.9 |
| Kusini Unguja | 392 | 46.9 | 13.5 | 1.6 |
| Mjini Magharib | 901 | 43.6 | 9.8 | 1.9 |
| Kaskazini Pemba | 446 | 43.5 | 13.4 | 1.6 |
| Kusini Pemba | 437 | 42.3 | 15.8 | 2.1 |
| **Wealth quintile** |  |  |  |  |
| Lowest | 530 | 44.5 | 13.8 | 2.5 |
| Second | 493 | 43.8 | 11.6 | 1.8 |
| Middle | 513 | 47.0 | 11.3 | 2.3 |
| Fourth | 538 | 45.5 | 12.3 | 0.9 |
| Highest | 482 | 41.9 | 11.0 | 1.5 |
| Total | 2556 | 41.9 | 11.0 | 1.5 |

**Table 3:**
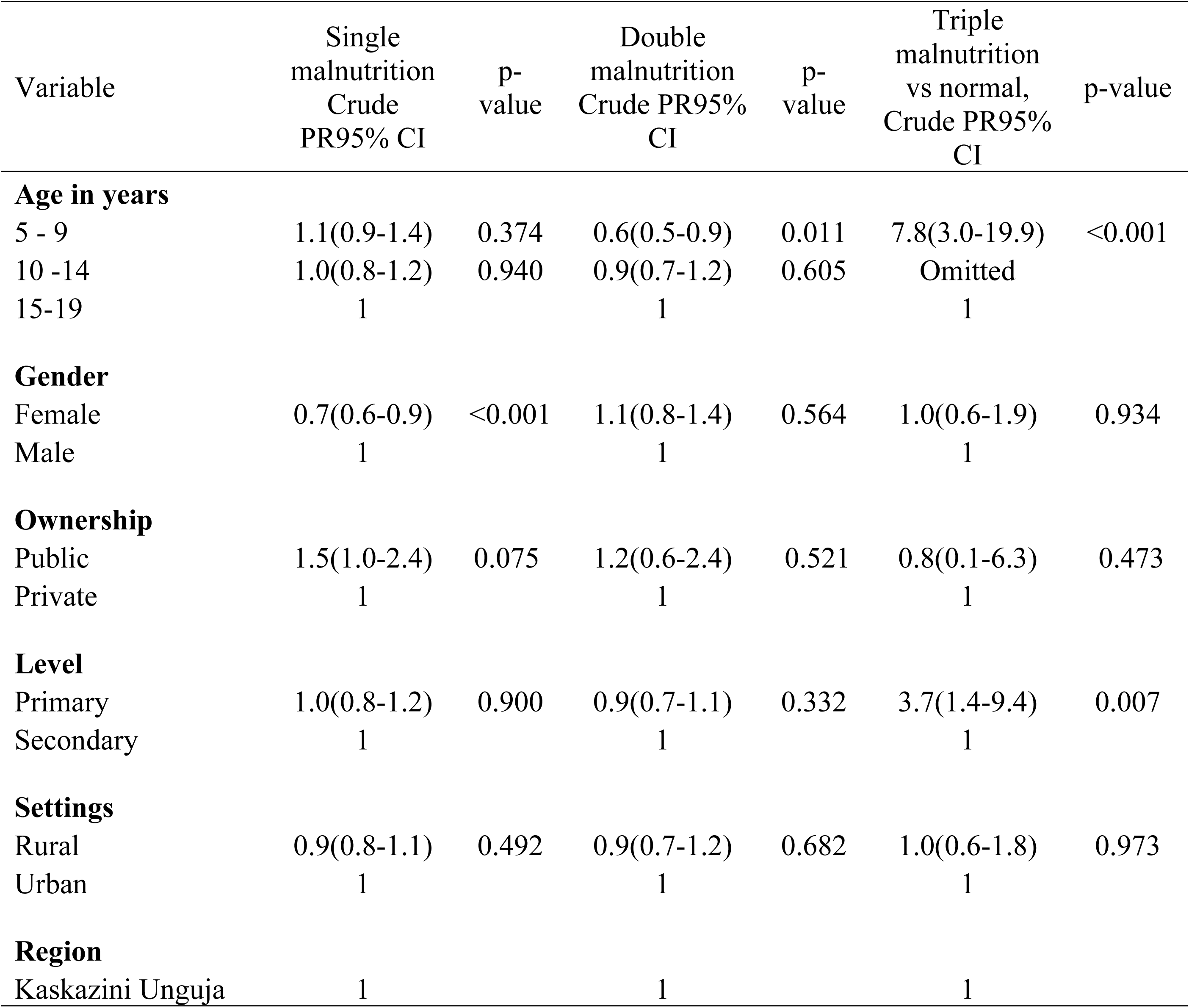
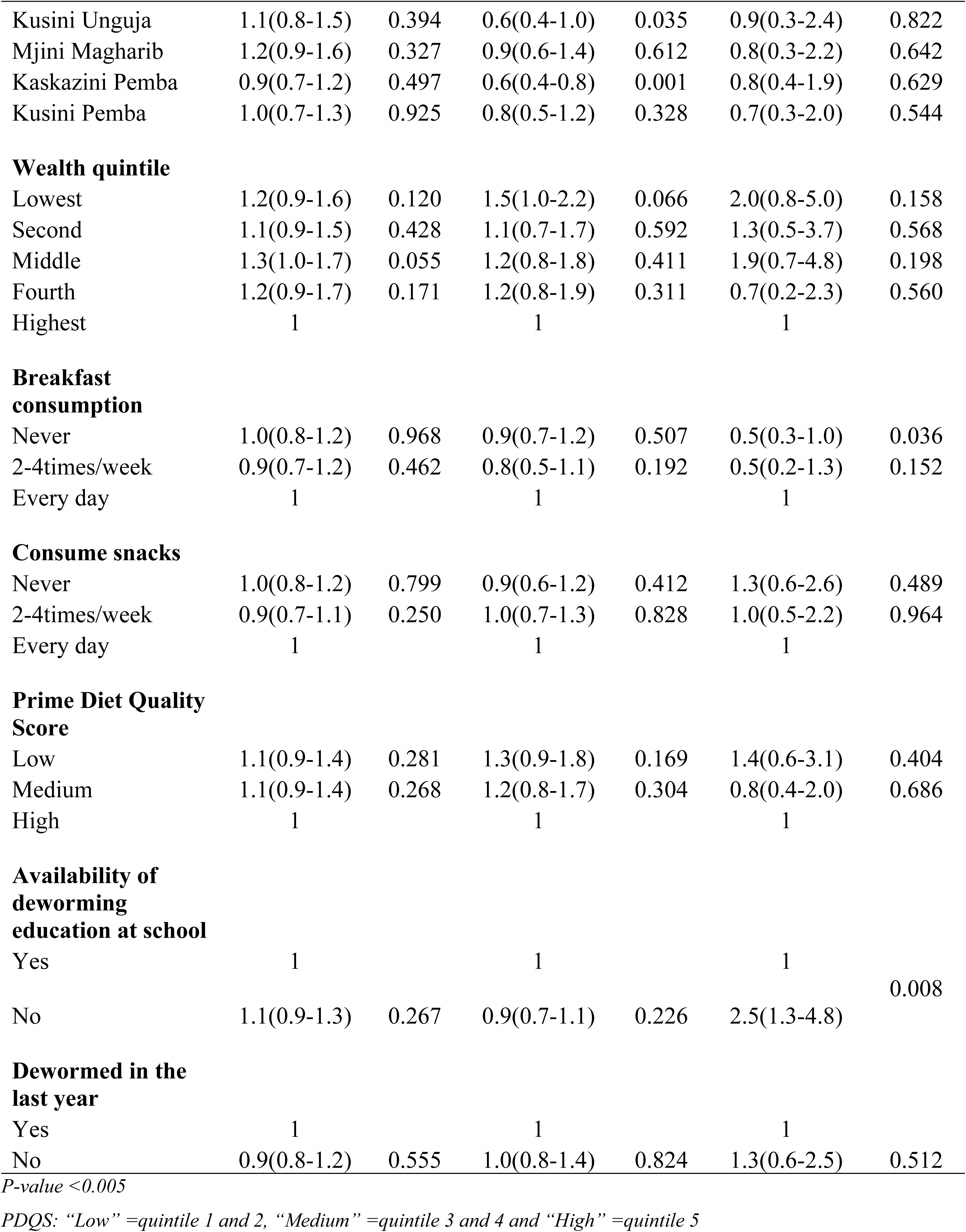
Determinants of malnutrition among primary and secondary school scholars in Zanzibar using unadjusted multinomial regression model (n=2556)

**Table 4:**
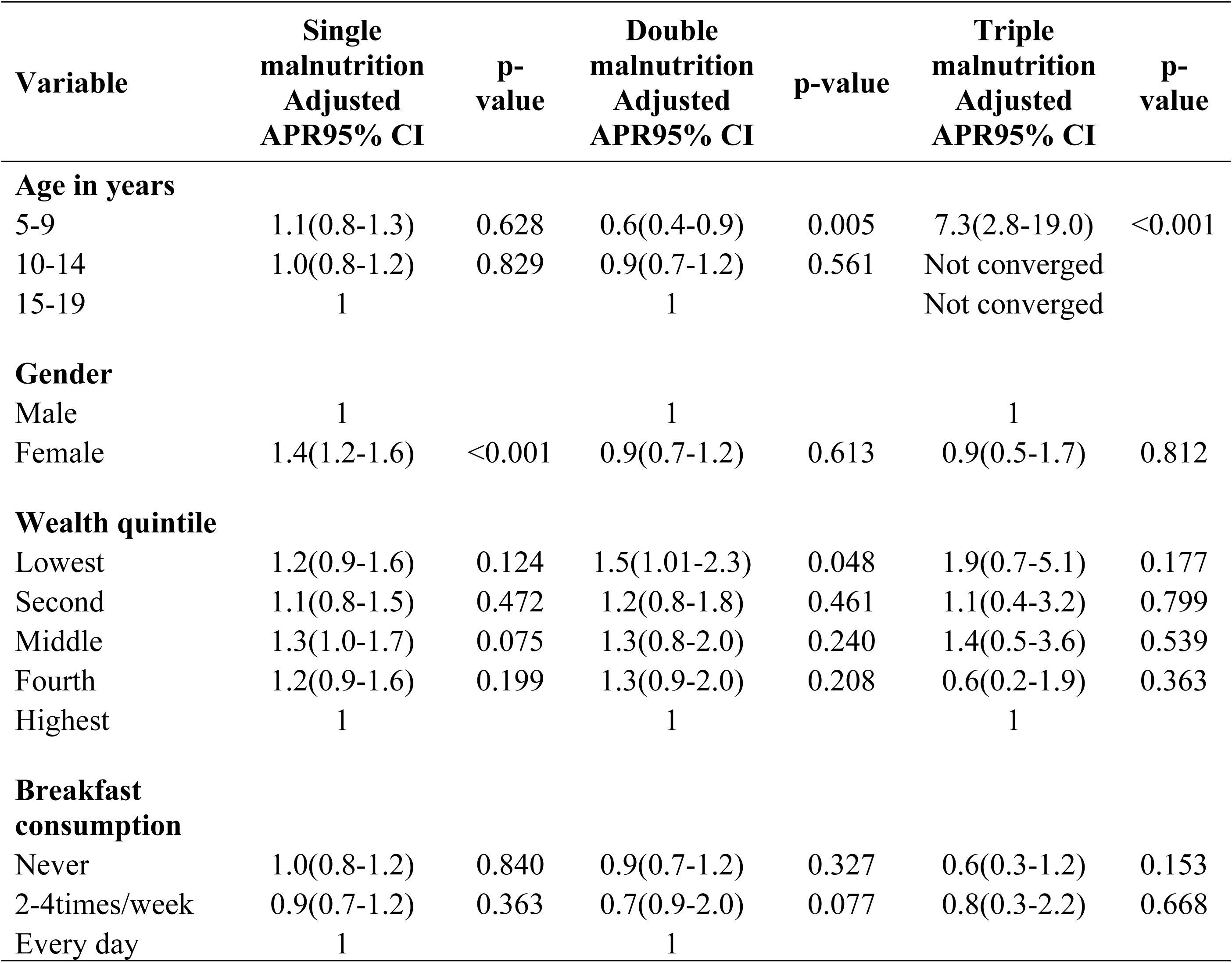
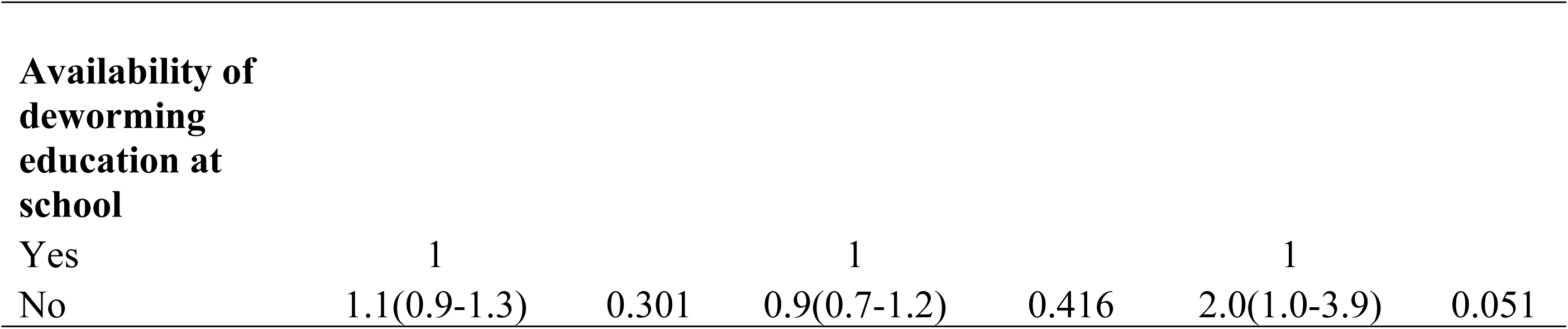
Determinants of malnutrition among primary and secondary school students in Zanzibar using adjusted multinomial regression model (n=2556)

| Variable | Single malnutrition<br>Adjusted APR95% CI | p-value | Double malnutrition<br>Adjusted APR95% CI | p-value | Triple malnutrition<br>Adjusted APR95% CI | p-value |
| --- | --- | --- | --- | --- | --- | --- |
| **Age in years** |  |  |  |  |  |  |
| 5-9 | 1.1(0.8-1.3) | 0.628 | 0.6(0.4-0.9) | 0.005 | 7.3(2.8-19.0) | <0.001 |
| 10-14 | 1.0(0.8-1.2) | 0.829 | 0.9(0.7-1.2) | 0.561 | Not converged |  |
| 15-19 | 1 |  | 1 |  | Not converged |  |
| **Gender** |  |  |  |  |  |  |
| Male | 1 |  | 1 |  | 1 |  |
| Female | 1.4(1.2-1.6) | <0.001 | 0.9(0.7-1.2) | 0.613 | 0.9(0.5-1.7) | 0.812 |
| **Wealth quintile** |  |  |  |  |  |  |
| Lowest | 1.2(0.9-1.6) | 0.124 | 1.5(1.01-2.3) | 0.048 | 1.9(0.7-5.1) | 0.177 |
| Second | 1.1(0.8-1.5) | 0.472 | 1.2(0.8-1.8) | 0.461 | 1.1(0.4-3.2) | 0.799 |
| Middle | 1.3(1.0-1.7) | 0.075 | 1.3(0.8-2.0) | 0.240 | 1.4(0.5-3.6) | 0.539 |
| Fourth | 1.2(0.9-1.6) | 0.199 | 1.3(0.9-2.0) | 0.208 | 0.6(0.2-1.9) | 0.363 |
| Highest | 1 |  | 1 |  | 1 |  |
| **Breakfast consumption** |  |  |  |  |  |  |
| Never | 1.0(0.8-1.2) | 0.840 | 0.9(0.7-1.2) | 0.327 | 0.6(0.3-1.2) | 0.153 |
| 2-4times/week | 0.9(0.7-1.2) | 0.363 | 0.7(0.9-2.0) | 0.077 | 0.8(0.3-2.2) | 0.668 |
| Every day | 1 |  | 1 |  |  |  |
| **Availability of deworming education at school** |  |  |  |  |  |  |
| Yes | 1 |  | 1 |  | 1 |  |
| No | 1.1(0.9-1.3) | 0.301 | 0.9(0.7-1.2) | 0.416 | 2.0(1.0-3.9) | 0.051 |

### Determinants of single and multiple (double & triple) malnutrition in Zanzibar using unadjusted multinomial regression model

The proportion of students and adolescents aged 5 to 19 years with single malnutrition in Zanzibar was 30% lower if the student is a girl (0.7 (95% CI, 0.6–0.9, p <0.001). Being 5 to 9 years and living in Kaskazini Pemba was a protective factor for having DBM. The prevalence of DBM was 40% lower if the child was 5 to 9 years 0.6 (95% CI, 0.5–0.9), p = 0.01), and if they were living in Kaskazini Pemba (0.6(95% CI, 0.4-0.8); p=0.001). The proportion of students with TBM was significantly very high among 5-9 year (7.8 (95% CI, 3.0–19.9, p < 0.001), and 3.7-fold greater if they were in a primary school (3.7(95% CI, 1.4-9.4); p=0.007). Double and single burden of malnutrition was associated with economic levels of the household where the child was living. While the proportion of students with DBM was 1.5-fold greater if a student was living in the lowest wealthy quantile (1.5(95%CI 1.0-2.2) students from middle wealth quantile had 1.3-fold greater chances of having single malnutrition compare to those from highest wealth quantiles (1.3(95%CI, 1.0-1.7); p=0.005)

### Determinants of single and multiple (double & triple) malnutrition in Zanzibar using adjusted multinomial regression model

In an adjusted multinomial regression model, the prevalence of DBM was 30% lower if the child was 5 to 9 years (0.7 (95% CI, 0.5–0.9), p = 0.02), and 1.5-fold greater if the child was living in a lowest wealth quantile family (1.5(95%CI, 1.01-2.3), p=0.04). In the contrary, the prevalence of single malnutrition was 1.4-fold greater if the student was a girl (1.4(95%CI,1.2-1.6); p=0.001), and 1.3-fold higher if the student was living in a middle wealth quantile family (1.3(95%CI, 1.0-1.7). Proportion of students with TBM was 2.0-fold greater if there were no school deworming education (2.0(95%CI, 1.0-3.9); p=0.05)

## DISCUSSION

This school-based cross-sectional study was conducted in Zanzibar to determine the prevalence and factors associated with the burden of malnutrition among students in Zanzibar. The study was conducted in recognition of the higher prevalence of overweight and obesity, especially among women of reproductive age, compared to the Tanzania mainland. Nevertheless, there is limited information on the burden of malnutrition among children and adolescents aged 5-19 years in Zanzibar despite rapid sociodemographic transitions, associated with urbanization, western dietary habits, and declining physical activities among school children in Zanzibar (4). To achieve our study objective, we used data from the National School Health and Nutrition Survey of 2022 with a focus on primary and secondary school children to generate data on anthropometric measurements complicated with the determination of hemoglobin levels.

The findings of this study can be categorized into three groups. The first group are the findings related to the nutritional status among children and adolescents in Zanzibar. Our study documented the prevalence of 45.7 per cent, 15.6 per cent, 13.9 per cent and 8.4 per cent for anemia, stunting, underweight and obesity. The overall prevalence of malnutrition was 58. 4 per cent. Looking at previous studies, school children in Zanzibar appear to have an unfavorable nutritional status than in some African countries and favorable to some. Our findings for example indicate a worsening trend of nutritional status in Zanzibar as compared to previous DHS data. Notably, the 2015-16 DHS data reported that 4 per cent of children in Zanzibar were underweight (6) which is unfavorable as compared to the present study but with favorable wasting prevalence (20-24 per cent) as compared to the findings of the present study. Furthermore, the stunting (15.6 per cent) and underweight (13.9 per cent) status of school children in Zanzibar appears to be slightly more favorable than children in public primary schools in Nairobi city of Kenya which were documented to be 24.5 per cent and 14.9 –per cent of wasting (7). Likewise, the prevalence of stunting and being underweight in Zanzibar appeared to be more favorable than children in Bangladesh documented to be 38 per cent and 40.5 per cent respectively (8). However, such favorability of nutritional status among school children in Zanzibar may be partly described by the sample size, age of students and the coverage of these studies. For instance, while our study gathered data from 2556 school children aged 5-19 years from private and public schools and rural and urban schools in all 11 districts of Zanzibar, the study by Mwaniki & Makokha (7) only examined the nutrition status and associated factors among 208 students aged 4–11years from four public primary schools in Dagoretti Division of Nairobi. Likewise, a study by Khanam & Haque (8) in Bangladesh focused on 400 children aged 5-10 years from only one district. This points to a suggestion that studies including both private and public schools in rural and urban contexts considering national coverage may unmask unfavorable nutritional status among school children and adolescents. Nevertheless, while the nutritional status among school children in Zanzibar may indicate favorable values as compared to some of the previous studies, these values are still alarming. This indicates a need for responsive interventions to improve nutritional status among school children in Zanzibar.

The second group of findings are related to the double Burden of Malnutrition (DBM) and the triple burden of Malnutrition (TBM). We defined DBM as the coexistence of both undernutrition and over-malnutrition and TBM as comprising three types of nutritional problems: undernutrition, overnutrition/obesity, and anemia). The WHO (9) recognizes DBM and TBM as among the serious forms of malnutrition that require immediate actionable interventions. Likewise, local data indicates that Tanzania is also experiencing the double burden of malnutrition, with 28 per cent of women and 4 per cent of children under five years suffering from overweight and obesity (6, 10). While most of the documented double burden of Malnutrition in Africa, particularly sub-Saharan Africa focuses on women and under five children (6,10–13), there are limited scholarly studies examining both the double and triple burden of malnutrition particularly among schoolchildren in Tanzania and other African countries. Our study findings indicated the prevalence of DBM and TBM of 12 per cent and 1.8 per cent among school children aged 5-19 years in Zanzibar respectively. While there are limited previous studies to which we can compare our findings, it is worth recognizing some that have mostly examined the triple burden of malnutrition among mother-child pairs. Notable mentions are the national family health survey in India indicating a TBM prevalence of 5.7 per cent (14) and a cross-sectional survey of low-income and middle-income countries documenting a TBM prevalence of 11.4 per cent (15). Of particular interest is a study by Modjadji & Madiba (16) in rural contexts of South Africa also focusing on mother-primary school children’s pairs with a TBM of 2.5 per cent (4 per cent overweight and one per cent obesity) among school children. Taken together, our study fills the gap for the DBM and TBM studies in Tanzania, the need for more context-specific studies is evident to inform evidence-based responsive interventions.

The third and final group are the findings related to the determinants of nutritional status, DBM and TBM. Our findings indicated that the prevalence of single malnutrition (stunting, underweight and obesity) and DBM was evenly distributed by age but was significantly higher among girls than boys. Schools in some areas of Pemba had a low prevalence of DBM. However, the burden of TBM was higher among 5 to 9 higher children with no gender variation in the proportion of school children with TBM. School children and adolescents in public and primary schools and schools with no deworming education had the highest prevalence of TBM. Contrastingly, there appear to be variations in predictors of nutritional status, BDM and TBM between our study and previous studies. For example, similar to our findings, girls were found to have more likelihood of being stunted and underweight in a study conducted in Bangladesh (8). In the cited study, the prevalence of stunting and underweight were 39 per cent and 45 per cent among girls whereas the prevalence was 36 per cent and 36 per cent among boys, respectively (8). However, contrary to our study, the likelihood of becoming malnourished was found to increase with the increased ages of the children, from the ages 5–6 years upward (8). Furthermore, while there were no gender variations of TBM among school children in our study, Boys were more likely to have TBM in a study conducted among mother-primary school children’s pairs in South Africa (16). Similarly, mothers’ socio-demographic characteristics such as age, educational status, cesarean section, birth size of baby, wealth status of a household, and place of residence were the most important correlates for the TBM among mother-child pairs in India (14) and in low and middle-income countries (15). Collectively, these findings point to a suggestion that more studies are needed to quantify the predictors of nutritional status, BDM and TBM among school children and adolescents to inform responsive interventions. This is because investment in school-going children and adolescents has been promoted as one of the key strategies in the realization of human potential for economic development (17).

This study is not without limitations. Although the findings address the research gap in Zanzibar and may be useful in policy formulation, the study is limited to information examining the predictors of poor nutritional status, DBM and TBM on the caregiver’s and community’s side. Further studies that tap into the insights of community members on the determinants of poor nutritional status, DBM and TBM are recommended for generating a comprehensive understanding of the persistent contributors of malnutrition among children in Zanzibar. Likewise, the study only focused on school children in Zanzibar. Future studies may seek to extend beyond schools to out-of-school children who may be at a disadvantage as compared to those attending schools.

## CONCLUSION

Our study unmasks novel findings related to the nutritional status, DBM and TBM among school children and adolescents aged 5-19 years in public and private schools in Zanzibar. The prevalence of anemia, stunting, underweight, obesity and DBM appears to be alarming. The prevalence of TBM is also high which underscore the need for targeted interventions. With most studies on DBM and TBM focusing on mother-child pairs, there is a need for more studies to generate more evidence on the predictors of nutritional status, BDM and TBM among school children and adolescents. Nevertheless, our findings indicate a need for the promotion of public health programs to create awareness about the harmful effects of sedentary lifestyles among schoolchildren. The findings also indicate the need for nutrition programs targeting undernutrition and anemia among school children and adolescents with a special focus on girls.

## Data Availability

Data will be shared with the authorization of the author

## Acknowledgement

The Ministry of Health (MoH) Zanzibar would like to express its gratitude and appreciation to all individuals, institutions, and partners who contributed to the implementation of this School Health and Nutrition Survey (SHNS) conducted in Zanzibar. In particular, the MoH acknowledges the contribution of partners from governmental agencies, the Office of the Chief Government Statistician (OCGS), the Ministry of Education and Vocational Training (MoEVT) together with the Tanzania Food and Nutrition Centre for their active participation and technical inputs toward this survey.

Special thanks go to the United Nations Children’s Fund (UNICEF) for their technical and financial support, and for seeing the requirement to adopt a similar study on Tanzania Mainland. The MoH would also like to thank all individual consultants, oversight organisations and partners for their roles in organising and preparing questionnaires, training of field staff, co-ordination of fieldwork, and support in developing this report, who include individuals from the Muhimbili University of Health and Allied Sciences (MUHAS), the National Institute for Medical Research (NIMR), and the Nelson Mandela University.

The MoH would also like to thank all the field supervisors, who oversaw the data collection exercise, and field data collectors who interviewed students. Moreover, the MoH would like to extend its appreciation to Data Entry Clerks, Regional and District teams (e.g., Malaria Focal Persons, laboratory technologists/technicians, School Health Programme Co-ordinators, Nutrition Officers, and drivers) for their participation during the survey. Furthermore, the MoH would like to thank all the students and school teachers for their readiness to participate in the survey. The MoH would also like to acknowledge the following teams of experts involved in the report preparation and finalisation:

Data analysis, Report writing and finalisation Team: Dr. Geofrey Mchau, Sauli Epimark, Shemsa Msellem, Hawa Msola, Fatma Abdallar, Dr Anna Mosses, Dr David Solomon and Asha Samini, Fatma Said, Adam Haji Ame, Heavenlight Ayubu, Stanlaus Erick Mollel, Salum Kassim Ally, Fahima Mohamed Issa, Sabina Raphael Daima, Kombo Mdachi Kombo, Hadija Hamisi Hamadi, Ahmad Hamza Mohamed, Aisha Mohamed Said, Asha Mahafodhi, and Prof. Esther Ngadaya.

